# Assessing spread risk of Wuhan novel coronavirus within and beyond China, January-April 2020: a travel network-based modelling study

**DOI:** 10.1101/2020.02.04.20020479

**Authors:** Shengjie Lai, Isaac I. Bogoch, Nick W Ruktanonchai, Alexander Watts, Xin Lu, Weizhong Yang, Hongjie Yu, Kamran Khan, Andrew J Tatem

## Abstract

**Background:** A novel coronavirus (2019-nCoV) emerged in Wuhan City, China, at the end of 2019 and has caused an outbreak of human-to-human transmission with a Public Health Emergency of International Concern declared by the World Health Organization on January 30, 2020.

**Aim:** We aimed to estimate the potential risk and geographic range of Wuhan novel coronavirus (2019-nCoV) spread within and beyond China from January through to April, 2020.

**Methods:** A series of domestic and international travel network-based connectivity and risk analyses were performed, by using de-identified and aggregated mobile phone data, air passenger itinerary data, and case reports.

**Results:** The cordon sanitaire of Wuhan is likely to have occurred during the latter stages of peak population numbers leaving the city before Lunar New Year (LNY), with travellers departing into neighbouring cities and other megacities in China. We estimated that 59,912 air passengers, of which 834 (95% UI: 478 - 1349) had 2019-nCoV infection, travelled from Wuhan to 382 cities outside of mainland China during the two weeks prior to Wuhan’s lockdown. The majority of these cities were in Asia, but major hubs in Europe, the US and Australia were also prominent, with strong correlation seen between predicted importation risks and reported cases. Because significant spread has already occurred, a large number of airline travellers (3.3 million under the scenario of 75% travel reduction from normal volumes) may be required to be screened at origin high-risk cities in China and destinations across the globe for the following three months of February to April, 2020 to effectively limit spread beyond its current extent.

**Conclusion:** Further spread of 2019-nCoV within China and international exportation is likely to occur. All countries, especially vulnerable regions, should be prepared for efforts to contain the 2019-nCoV infection.

## Introduction

In December 2019, a cluster of patients with pneumonia of unknown cause were reported in the city of Wuhan, Hubei Province, China, and epidemiologically linked to a seafood wholesale market [1, 2]. It has been determined that the pathogen causing the viral pneumonia among affected individuals is a new coronavirus (2019-nCoV) [1, 3]. The pathogen exhibits high human-to-human transmissibility and has spread rapidly within and beyond Wuhan city [4, 5]. On January 30^th^, 2020, World Health Organization (WHO) declared the 2019-nCoV outbreak a Public Health Emergency of International Concern [6].

Wuhan is central China’s transportation hub with a population of 11 million residents and a large number of higher-education students (∼1.3 million in 89 universities and colleges), a particularly mobile population [7]. Beyond these factors, viral spread was likely exacerbated further by the surge in domestic and international travel during the 40-day Lunar New Year (LNY) celebrations (from January 10^th^, 2020 to February 18^th^, 2020) – the largest annual human migration in the world, comprised of hundreds of millions of people travelling across the country. As of February 4^th^, 2020, China has reported 20,530 confirmed cases and 23,314 suspected cases with 2019-nCoV infections [8]. Of the confirmed cases, 2788 are severe and 426 people have died. Most cases were reported from Wuhan and other cities in Hubei Province, and all provinces have confirmed cases imported from Wuhan and secondary transmission has been reported in some provinces. Additionally, there were 153 cases reported in 23 countries outside of China, with most having a travel history involving Wuhan [6].

The potential pathway from this local outbreak in Wuhan to a pandemic might involve four steps: i) local transmission in Wuhan (primary city of epidemic); ii) spread from Wuhan to other cities within and beyond China via infected travellers, causing sporadic secondary transmission in these cities (secondary city of epidemic), iii) further spreading from secondary cities with local transmission to other tertiary cities in China and international cities via returning travellers after the LNY holiday; iv) onward transmission across multiple countries and leading to a pandemic. To interrupt the spread, a cordon sanitaire of Wuhan and surrounding cities in Hubei Province has been in place since January 23^rd^, 2020, just two days before the LNY’s Day. However, significant numbers of people had likely already travelled back to their hometowns for the holiday by this time. According to Wuhan authorities, it is likely that more than five million residents had already left the city before the lockdown, but where they went and how high the risk of spreading the virus remains an open question [9].

Here we conducted a travel network-based analysis to explore patterns of domestic and international population movements from high-risk cities in China, and provide preliminary estimates of the potential risk of 2019-nCoV spreading across and beyond the country. Given the current epidemic and limited understanding of the epidemiology of this disease, our findings on travel patterns from historical data can help contribute to tailoring public health interventions.

## Methods

To identify the areas that are most vulnerable to virus importation, we performed and integrated a series of analyses, by using de-identified and aggregated mobile phone-based population movement data, air passenger itinerary data, and case reports. We defined the potential risk and geographic range of 2019-nCoV virus spread across three scenarios: 1) from the primary city (Wuhan) into other cities in mainland China (31 provincial regions), 2) from high-risk secondary cities into other cities across China, and 3) from high-risk cities in mainland China into cities in other countries or regions during the LNY holiday and the following three months. We also estimated the number of airline travellers likely needing to be quarantined or screened to capture travellers potentially exposed to 2019-nCoV in high-risk cities of mainland China.

### Spread risk and destinations of 2019-nCoV from Wuhan

To define daily patterns and the connectivity of population movements at county and prefecture (city) level across mainland China during the LNY holiday and the following three months, we used the aggregated and de-identified daily flow of the users of Baidu, the largest Chinese search engine [10]. Baidu offers location-based service (LBS), based on the global positioning system (GPS), IP address, location of signalling towers and WIFI, for online searching, mapping, and a large variety of apps and software for mobile devices. These data have been used to visualize population migration around Chinese New Year [10]. Two Baidu datasets were used in this study. The first one covers daily movement data at county level from December 1^st^, 2013 to April 30^th^, 2014, as described elsewhere [11]. We calculated relative netflow following the equation below to extract daily patterns during LNY holiday at county level, with the population in 2014 obtained from the Chinese Bureau of Statistics [12].

### Relative netflow = (inflow – outflow) / population of each county

The second dataset is a more recent daily movement matrix at the city level based on data from Baidu’s search app from January 1^st^, 2015 to April 30^th^, 2015. The last recorded locations of a user (or device) for each day were compared, and if the location changed, then we counted the user (or device) as someone who had moved from one city to another city. To understand the spread risk of 2019-nCoV from Wuhan into other cities via domestic population movement, we aggregated daily outflows of people from Wuhan to other cities across mainland China for the two weeks (the quarantine period of the virus) prior to the cordon sanitaire of Wuhan. As the travel ban took place on January 23^rd^, 2020, just 2 days before LNY’s day, and given that LNY’s day in 2015 was February 19^th^, we took February 17^th^, 2015 as a reference of the lockdown day in our dataset. Using the second dataset, then the risk of importation for each destination city or province was preliminarily defined as the percentage of travellers received by each city or province out of the total volume of travellers leaving Wuhan during the two weeks before the city’s lockdown.

### Spread risk from high-risk secondary cities in mainland China

These secondary cities may have a high risk of community-level transmission through the introduction of infected travellers from Wuhan, and then spread the virus to other tertiary cities by returning population movements after the LNY holiday, causing an even wider spread of the virus. As most of the cities in Hubei province have implemented the same travel controls as Wuhan before LNY, we defined the high-risk secondary cities outside of Hubei province as the cities within top 30 ranked cities (Supplementary Table S1) with the highest risk of importation from Wuhan defined above. Based on the 2015 Baidu dataset on population movement, the risk of spreading the virus from high-risk secondary cities to tertiary cities was preliminarily calculated as the averaged percentage of travellers received by each tertiary city out of the total volume of travellers leaving each high-risk secondary city during the four weeks following LNY’s Day. We chose a period of four weeks because the returning flow of LNY’s population movement, *Chunyun*, generally lasts four weeks.

### Destinations of the virus spreading beyond mainland China

To define the connectivity and risk of 2019n-CoV spreading from Wuhan and high-risk secondary cities defined above, into the cities beyond mainland China, we obtained aggregated itinerary data from the International Air Travel Association (IATA) [13]. IATA data accounts for approximately 90% of passenger travel itineraries on commercial flights, and these data represent direct origin (Wuhan) to destination trips, and indirect trips that originated in Wuhan, but had connecting flights to a final destination. We quantified monthly volumes of airline travellers departing Wuhan and high-risk secondary cities from February 1^st^, 2018, through April 30^th^, 2018. With the assumption that the population movements around the LNY holiday in 2020 was consistent with the pattern in 2018, all final destinations were ranked by volumes of airline travellers, and the relative risk of importation was defined as the percentage of airline travellers received by each destination city out of the total volume of travellers leaving high-risk cities in China.

We also estimated the number of airline travellers that may have needed to be kept in quarantine from Wuhan during the two weeks prior to the city’s travel ban. The LNY’s day in 2018 was on February 16 and the lockdown of Wuhan happened two days before LNY’s day, corresponding to the date of February 14^th^, 2018. We therefore defined the number of travellers needing to be quarantined as half of the volume of airline travellers from Wuhan in February, 2018, representing the 2-week total number of travellers for the first half of February. We then estimated the number of infections and its 95% uncertainty interval (UI) in these airline travellers from Wuhan, based on a binomial distribution and the proportion of 2019-nCoV infections in the citizens evacuated from Wuhan reported by Singapore, South Korea, Japan, and Germany, as of January 31^st^, 2020 [6, 14-17].

Additionally, to capture travellers potentially exposed to virus, we also estimated the volume of airline travellers that would be required to be screened at origin high-risk cities in China and destinations across the globe for the following three months of February to April. Considering air traffic flows from China have changed due to the airline flight cancellations, travel restrictions imposed by countries or regions, or changes in travel behaviours, we calculated the volume of travellers using different scenarios of reductions (50%, 75%, and 90%) in total passenger volumes.

### Validation

As we used historial data to predict travel patterns in 2020, to ensure that the seasonal patterns observed over LNY and holidays more generally are consistent over multiple years and countries, we collated country-level domestic and international passenger statistics for air travel from 2010 to 2018, and compared these against a comprehensive time series of public and school holidays across the world during the same period (Supplementary Note). Additionally, we also compared the spatial patterns of the risks of Chinese cities importing the virus from Wuhan, estimated by the population movement data in 2014 and 2015, and more recent data on the top 50 ranked origin and destination cities in January 2020, available from the Baidu Migration site (https://qianxi.baidu.com/). To futher validate our results, we also compared the importation risk estimated in this study with the number of reported imported cases from Wuhan to other provinces in mainland China, as of January 25^th^, 2020, and the number of imported cases reported by other countries or regions, as of February 3^rd^, 2020 [6]. The distribution between days of travelling from Wuhan, illness onset, first medical visit, and hospitalization of imported cases, as of January 25^th^, 2020, were also analysed. These case data were collated from the websites of WHO, national and local health authorities or new agencies within and beyond China (Supplementary Note). R version 3.6.1 (R Foundation for Statistical Computing, Vienna, Austria) was used to perform data analyses.

## Results

### Risk and destinations of virus spread within mainland China

Significant migratory flows occurred in opposite directions before and after LNY’s Day. In Wuhan City, mass movements of people began about three weeks prior to LNY, with the first peak of population leaving the city before the start of the winter holiday for universities, especially in the three counties that contain many universities and students (Figure 1). Although a cordon sanitaire of Wuhan and some cities in Hubei Province has been in place since January 23^rd^, 2020, the timing of this may have occurred during the latter stages of peak population numbers leaving Wuhan, as another peak of movements out of the city was seen 2 days before LNY’s day.

**Figure 1:**
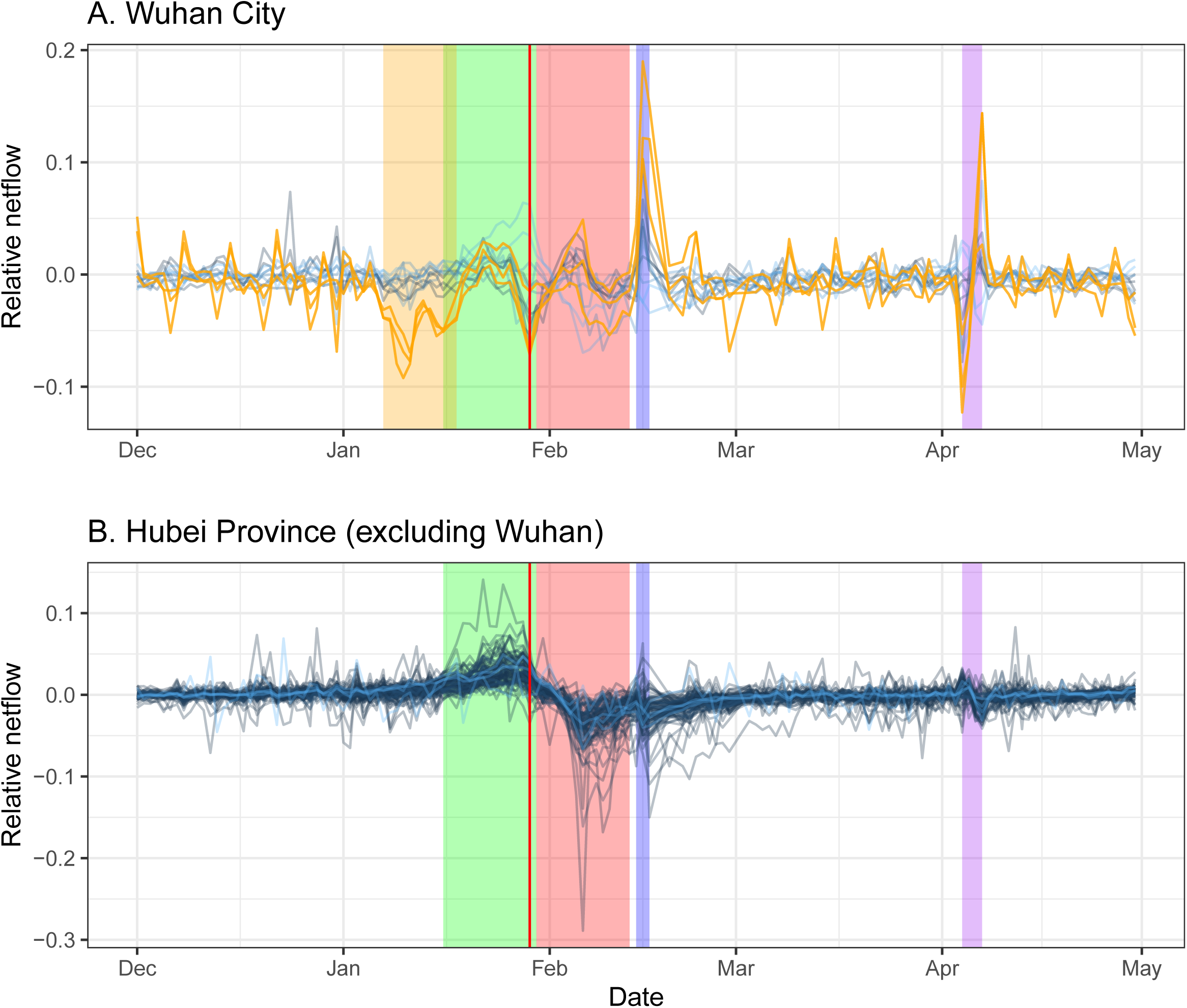
Patterns of daily human movement by county in Wuhan City and Hubei Province across five months. **(A)** Wuhan City. **(B)** Hubei province (excluding Wuhan). Each blue line reprents the netflow of population movement in each county. Yellow lines reprent the netflow of population movement of three counties (Wuchang, Hongshan, and Jiangxia) with more universities or collegues in Wuhan. Vertical red line shows the day of cordon sanitaire in place in cities of Hubei. Shadow colours: yellow – 2 weeks before the start of winter break of universities in Wuhan; green - 2 weeks before LNY’s Day; red - 2 weeks since LNY’s Day; blue - Lantern Festival and weekend; purple - Tomb Sweeping holiday and weekend. Relative netflow = (Inflow – Outflow)/population, based on the population movement data in 2013-2014 obtained from Baidu, Inc.

We found that a large number of travellers were likely departing Wuhan into neighbouring cities and other megacities in China before Wuhan’s lockdown (Supplementary Tables S1). Other cities in Hubei Province received huge amounts of people during the two weeks before LNY and showed a decreasing population since LNY, following a peak of outflow at the end of the LNY holiday (Figure 1b and Supplementary Table S2). If Wuhan’s lockdown had not have been undertaken, our analyses suggest the main destination cities of population outflow since LNY would have been similar to the situation two weeks prior to LNY (Supplementary Table S3).

In terms of the initial importation risk of virus for each city during the two weeks before Wuhan’s lockdown, nearly all other cities in Hubei Province were estimated to be high-risk areas (Figure 2a). Other places with high risks were Beijing, Shanghai, Guangzhou and other large cities. In terms of the provincial level, the risks were high in Guangdong and Hunan, followed by Henan and Zhejiang (Figure 2b). There was a significant correlation (r-squared = 0.59, p < 0.001) between the number of imported cases and the risk of importation estimated from traveller numbers from Wuhan within the two weeks before LNY’s Day (Figure 3a). Further, a high proportion of cases travelled with symptoms at the early stage of the outbreak, and the lag from illness onset to hospitalization decreased from a median of 6 days (Interquartile range: 4-7 days) in the first half of January 2020 to 3 days (1-5 days) in the second half (Figure 4).

**Figure 2:**
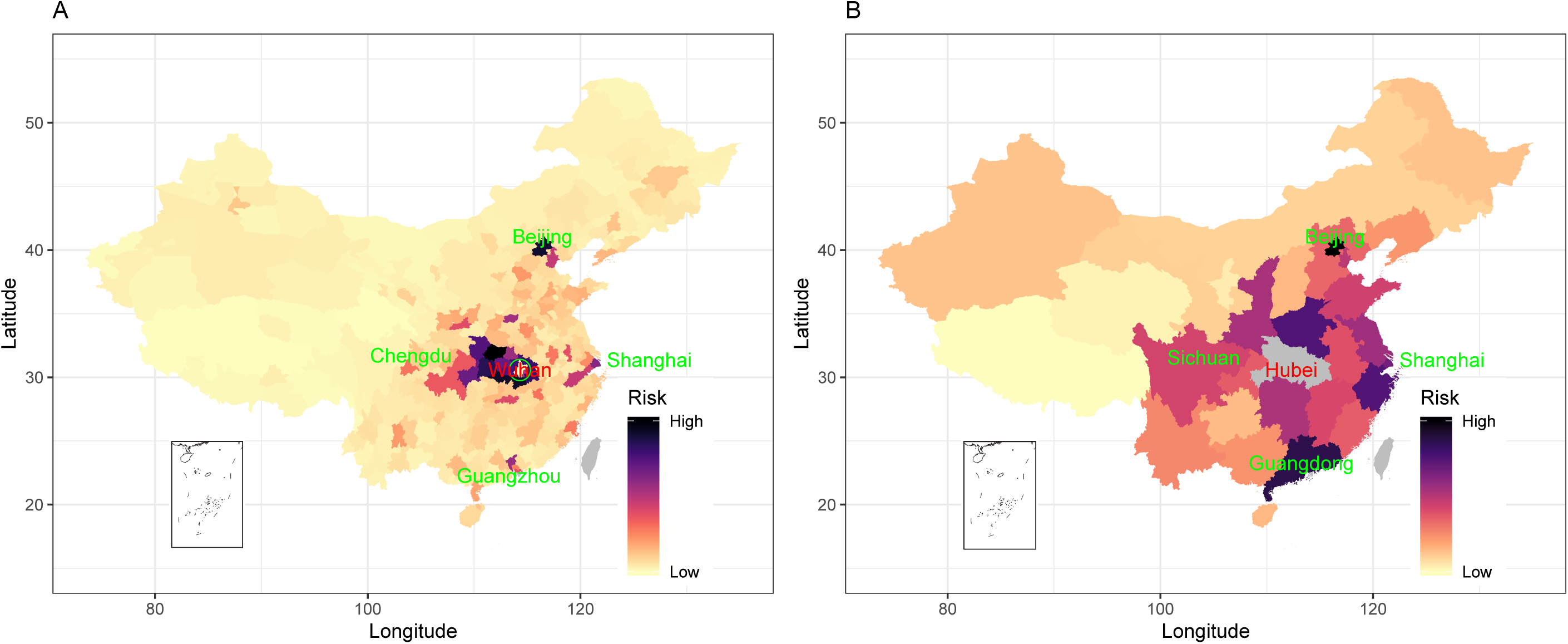
Risk of cities and provinces in mainland China receiving travellers with 2019-nCoV infections from Wuhan during the two weeks before the city’s lockdown. **(A)** at city level. **(B)** at provincial level (excluding Hubei). The risk of importation for each destination city or province was preliminarily defined as the percentage of travellers received by each city or province out of the total volume of travellers leaving Wuhan during the two weeks before the city’s lockdown.

**Figure 3:**
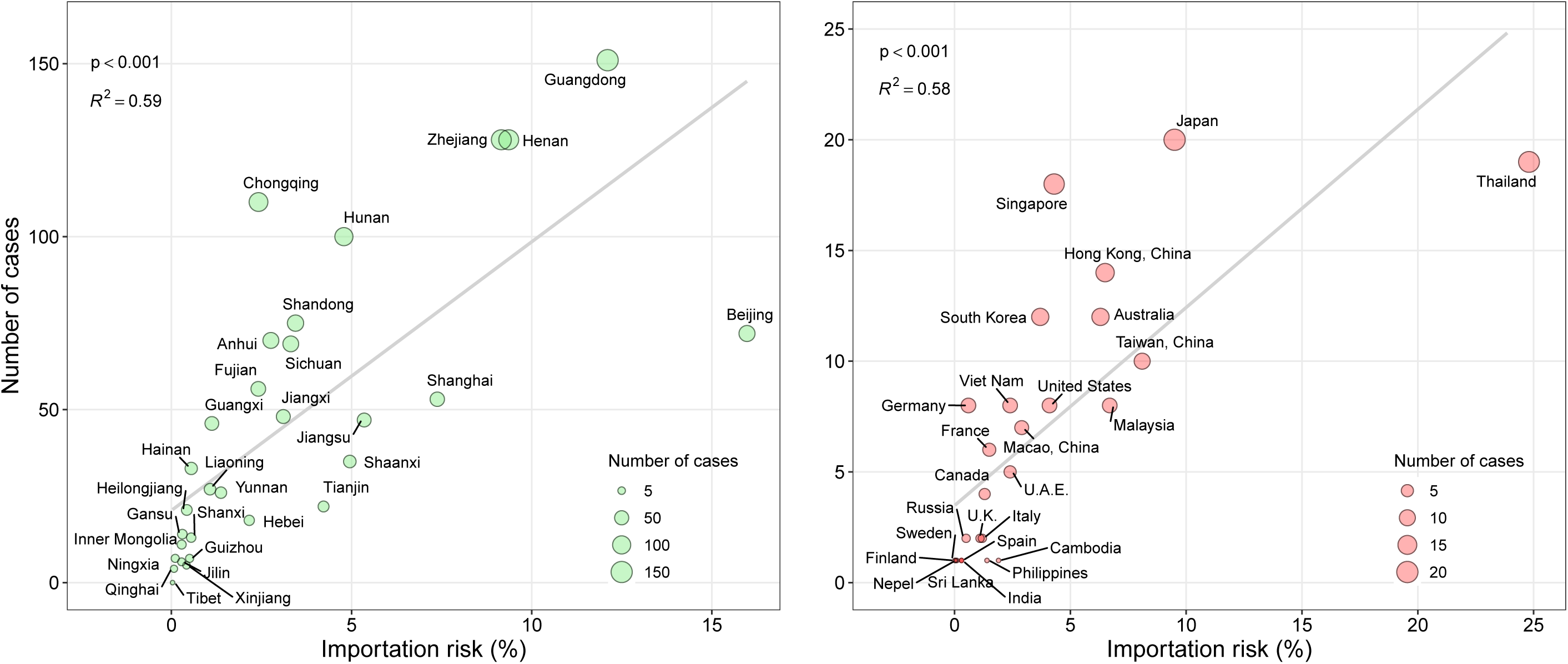
Correlation between the number of cases reported and the risk of importation via travel. **(A)** Number of imported cases reported by each province (excluding Hubei), as of January 25^th^, 2020, versus the risk of importation from Wuhan. The risk of importation for each province was preliminarily defined as the percentage of travellers received by each province out of the total volume of travellers leaving Wuhan during the two weeks before the city’s lockdown. **(B)** Number of imported cases reported by each country or region, as of February 3^rd^, 2020, versus the risk of importation from Wuhan. The risk of importation for each country or region was preliminarily defined as the percentage of travellers received by each destination out of the total volume of airline travellers leaving Wuhan from February to April 2018. Grey lines represent linear regression of importation risk against the number of cases reported, with R-squared and p-values are indicated on the graphs.

**Figure 4:**
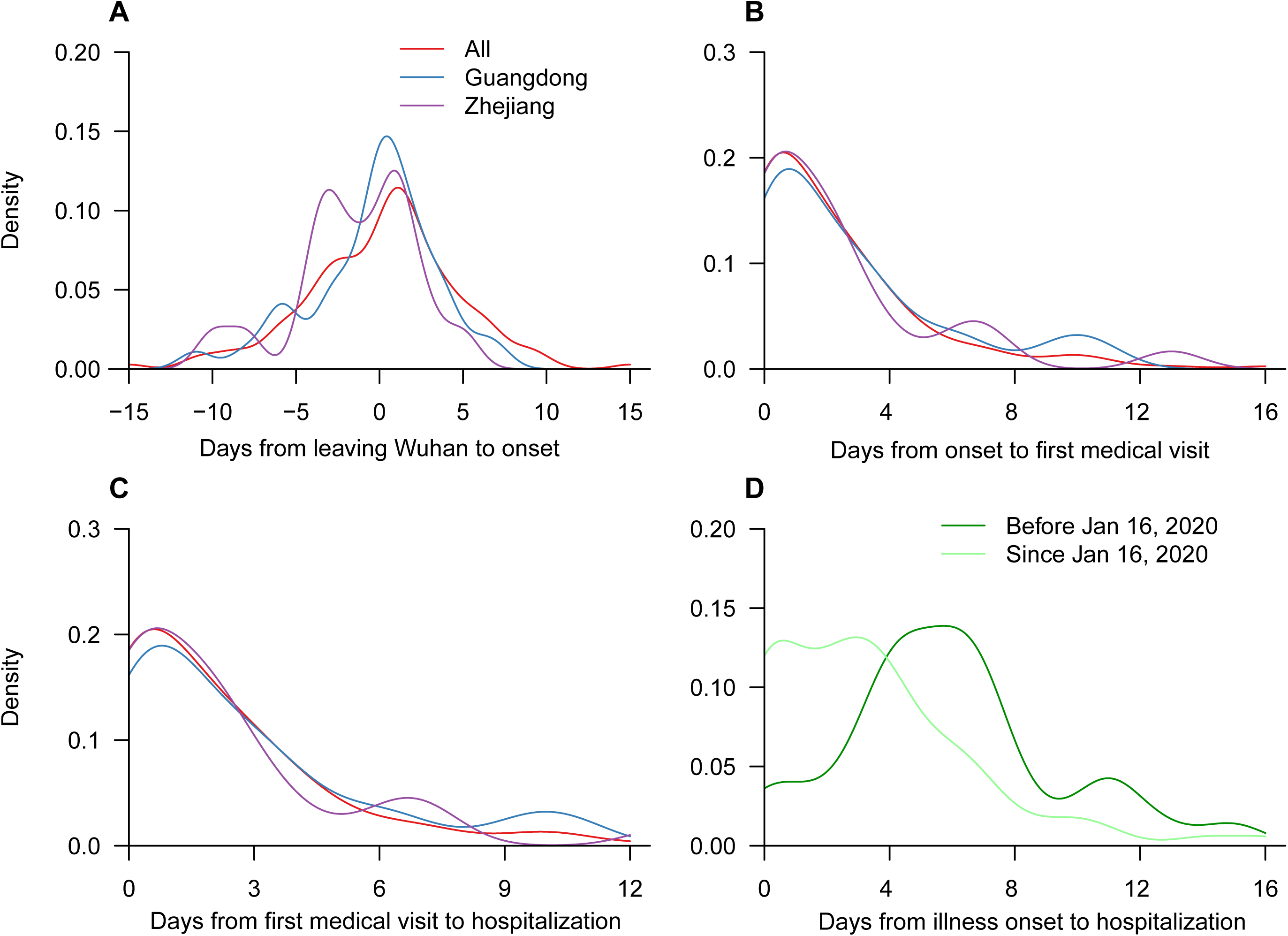
Time distributions of cases imported from Wuhan into other cities in China before the city’s lockdown on January 23^rd^, 2020. **(A)** Time difference from leaving Wuhan to illness onset (N=145). The negative days means onset of illness prior to travelling. **(B)** Time difference from illness onset to first medical visit (N=164). **(C)** Time difference from first medical visit to hospitalization (N=164). **(D)** Days from illness onset to hospitalization during the first half (N=67) and second half (N=80) of January, 2020, respectively. A total of 164 cases with available data as of January 25th, 2020, were included.

According to our definition outlined in the methods, the 17 high-risk secondary cities outside of Hubei Province were identified as: Beijing, Shanghai, Guangzhou, Zhengzhou, Tianjin, Hangzhou, Jiaxing, Changsha, Xi’an, Nanjing, Shenzhen, Chongqing, Nanchang, Chengdu, Hefei, Fuzhou, and Dongguan (Figure 5). Should community-level outbreaks occur in these cities, they could contribute to further spread of infection to other highly connected cities within China via movement after the holiday (Supplementary Figures S1 and S2).

**Figure 5:**
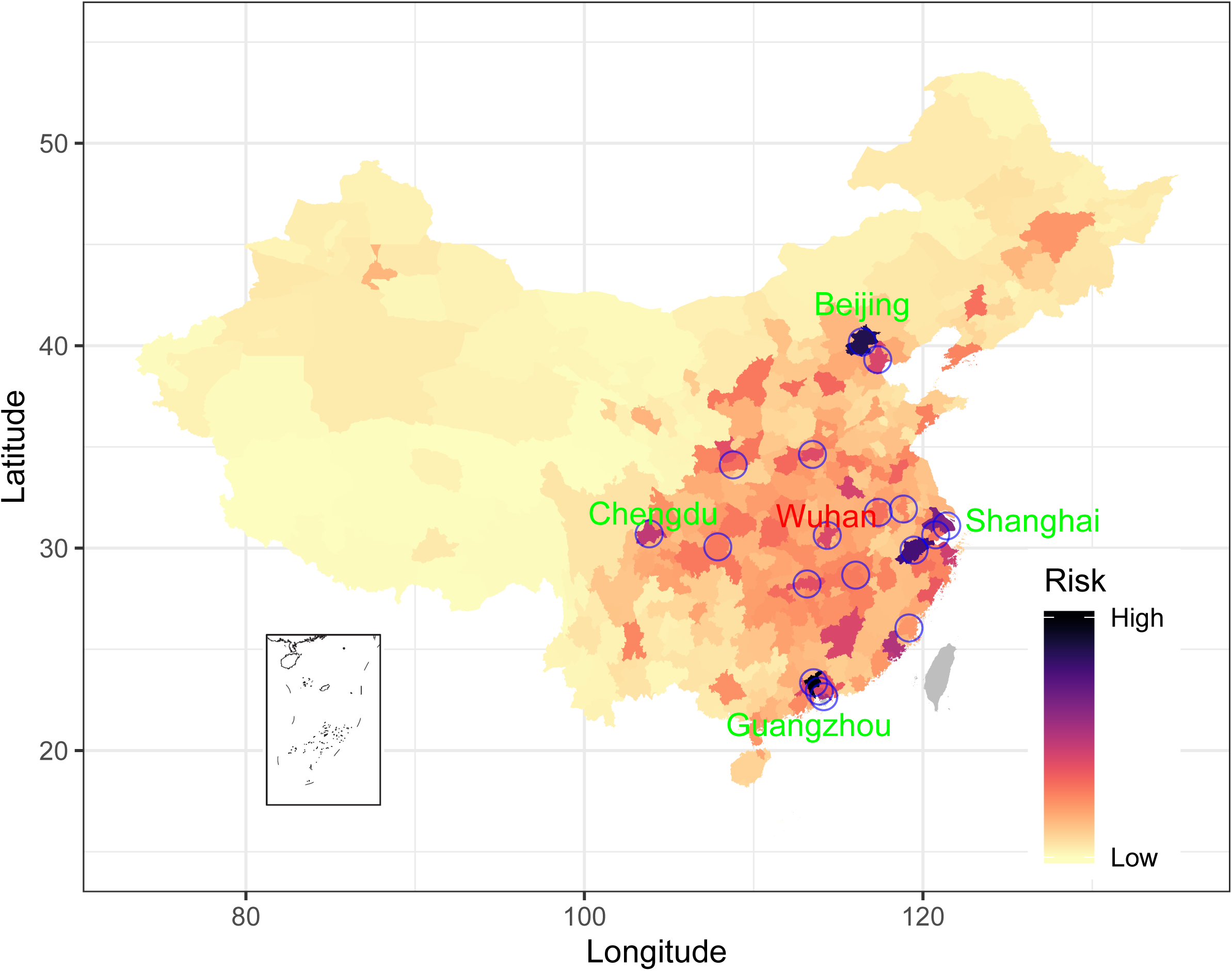
Risk of cities in mainland China receiving travellers from high-risk cities (blue circles) with 2019-nCoV infections or importations during the next four weeks since LNY’s Day. The risk of importation at city level was preliminarily defined as the averaged percentage of travellers received by each city out of the total volume of travellers leaving each high-risk city, based on the population movement data in 2015 obtained from Baidu, Inc. The high-risk cities include Wuhan in Hubei province and 17 cities (Beijing, Shanghai, Guangzhou, Zhengzhou, Tianjin, Hangzhou, Jiaxing, Changsha, Xi’an, Nanjing, Shenzhen, Chongqing, Nanchang, Chengdu, Hefei, Fuzhou, and Dongguan) in other provinces receiving high volume of travellers from Wuhan during the two weeks before the city’s lockdown on January 23^rd^, 2020.

### International spread risk and destinations

Based on historical air travel data, the connectivity and spread risk between high-risk cities in mainland China and cities in other countries or regions was defined for the three months around the LNY holiday (Supplementary Figure S3 and Tables S4 and S5). Bangkok, Hong Kong, and Taipei ranked in the top three, followed by Seoul, Tokyo and Singapore. The main destinations were presented by region in the supplemental materials (Supplementary Figures S3-S9 and Table S4-S7). During the two weeks before Wuhan’s lockdown, there were an estimated total 59,912 airline travellers from Wuhan that may have needed to be kept in quarantine at the 382 destinations outside of mainland China (Supplementary Table S8). Thailand, Japan, and Taiwan ranked in the top three, followed by Malaysia, Hong Kong, and Singapore. Based on an overall infection rate of 1.39% (16/1149; 95% UI: 0.80% - 2.25%) in citizens evacuated from Wuhan before February 1^st^, 2020, reported by Singapore (1.08%, 1/92), South Korea (1.36%, 5/368), Japan (1.42%, 8/565), and Germany (1.61%, 2/124), we made preliminary estimates of a total of 834 (95% UI: 478 - 1349) airline travellers that may have been infected with 2019-nCoV from Wuhan two weeks prior to the city’s lockdown. If adjusted by the estimated doubling time of the virus transmission from the lockdown to the evaualtion, 297 (170, 480) airline travellers may have been infected (Supplementary Table S8). As of February 3^rd^, 2020, a significant correlation (r-squared = 0.58, p < 0.001) was seen between the number of imported cases reported in those countries or regions and the risk of importation via travellers defined in our study (Figure 3b).

Because significant intranational and international spread from Wuhan has already occurred, a very large number of airline travellers (6.5 million under the scenario of 50% travel reduction as usual, 3.3 million under 75% reduction, and 1.3 million under 90% reduction, respectively) would be required to be screened at origin high-risk cities in China (Figure 5) and destinations across the globe for the following three months of February to April, 2020 (Table 1 and Supplementary Tables S9 and S10), to ensure that all travellers from high risk cities were covered.

Addtionally, based on monthly air passenger travel statistics and time series of public and school holidays in 91 countries from 2010 to 2018 (Supplementary Note), we found the seasonal pattens of domestic and international population movements across years were highly consistent with the timing and duration of public and school holidays in different countries (Supplementary Figure S10 and S11). Moreover, we also found similar spatial patterns in the risks of Chinese cities importing the virus from Wuhan via population movements estimated by both the Baidu data in 2014 and 2015, and the more recent data covering the top 50 ranked destinations in 2020, (Figure 1a and Supplementary Figure S12), highlighting the value of using historical data to rapidly assess present day risks.

**Table 1.** Top 30 ranked cities across the globe receiving airline travellers from 18 high-risk cities (Figure 5) in mainland China from February to April, representing three-month air traffic after LNY’s holiday with travel banned from Wuhan and 75% reduction of travel from othe cities.

| Rank | Top 30 countries or regions |  | Top 30 cities |  |  |
| --- | --- | --- | --- | --- | --- |
|  | Countries/regions | Volume (%)* | City | Countries/regions | Volume (%)* |
| 1 | Thailand | 485.6 (14.5) | Bangkok | Thailand | 254.2 (7.7) |
| 2 | Japan | 382.8 (11.5) | Hong Kong | Hong Kong, China | 244.4 (7.4) |
| 3 | Hong Kong, China | 244.4 (7.4) | Taipei | Taiwan, China | 209.3 (6.4) |
| 4 | Taiwan, China | 237.7 (7.3) | Seoul | South Korea | 185.8 (5.7) |
| 5 | South Korea | 230.2 (7.1) | Tokyo | Japan | 173.4 (5.3) |
| 6 | United States | 189.1 (5.1) | Singapore | Singapore | 138.6 (4.2) |
| 7 | Malaysia | 150.6 (4.4) | Phuket | Thailand | 118.8 (3.6) |
| 8 | Singapore | 138.6 (4.2) | Osaka | Japan | 107.0 (3.3) |
| 9 | Viet Nam | 115.6 (3.5) | Kuala Lumpur | Malaysia | 92.2 (2.8) |
| 10 | Australia | 109.8 (3.2) | Macau | Macau, China | 62.3 (1.9) |
| 11 | Indonesia | 99.6 (2.8) | Denpasar Bali | Indonesia | 53.1 (1.6) |
| 12 | Cambodia | 64.3 (1.9) | Sydney | Australia | 49.9 (1.5) |
| 13 | Macau, China | 62.3 (1.9) | Chiang Mai | Thailand | 37.8 (1.2) |
| 14 | Germany | 58.0 (1.8) | Los Angeles | United States | 38.0 (1.2) |
| 15 | Philippines | 61.6 (1.8) | Melbourne | Australia | 37.3 (1.1) |
| 16 | United Kingdom | 46.7 (1.4) | Nagoya | Japan | 34.6 (1.1) |
| 17 | Canada | 50.7 (1.4) | London | United Kingdom | 34.9 (1.1) |
| 18 | Italy | 36.9 (1.1) | New York | United States | 36.0 (1.1) |
| 19 | U.A.E | 38.7 (1.1) | Ho Chi Minh City | Viet Nam | 34.6 (1.1) |
| 20 | Russia | 36.9 (1.0) | Nha Trang | Viet Nam | 35.7 (1.1) |
| 21 | France | 32.9 (0.9) | Dubai | U.A.E | 34.4 (1.0) |
| 22 | India | 26.0 (0.8) | Phnom Penh | Cambodia | 31.2 (0.9) |
| 23 | New Zealand | 29.5 (0.8) | Siem Reap | Cambodia | 30.2 (0.9) |
| 24 | Spain | 26.1 (0.7) | Paris | France | 28.5 (0.9) |
| 25 | Egypt | 14.3 (0.4) | Kota Kinabalu | Malaysia | 29.6 (0.9) |
| 26 | Maldives | 12.2 (0.4) | Manila | Philippines | 29.5 (0.9) |
| 27 | Sri Lanka | 13.8 (0.4) | Krabi | Thailand | 29.8 (0.9) |
| 28 | Turkey | 16.5 (0.4) | Frankfurt | Germany | 25.5 (0.8) |
| 29 | Laos | 8.6 (0.3) | Jakarta | Indonesia | 27.8 (0.8) |
| 30 | Myanmar | 10.4 (0.3) | Kaohsiung | Taiwan, China | 24.8 (0.8) |
|  | Other | 254.6 (10.2) | Other |  | 1015.8 (30.8) |
|  | Total | 3285 (100) | Total |  | 3285 (100) |

\* In thousand. Based on air travel data from February to April 2018, obtained from the International Air Travel Association (IATA).

## Discussion

Mobile phone-based population movement data and air passenger itinerary data have been widely used to quantify the connectivity and transmission risk of pathogens via domestic and international human travel [18-23]. Given the rapidly growing number of confirmed 2019-nCoV infections, increasing evidence of human-to-human transmission within and beyond China [4, 24], and our limited understanding of this novel virus [25, 26], the findings here from travel patterns in historical data and spread risk estimation can help guide public health preparedness and intervention design across the world [27].

In terms of domestic connectivity and risk, the high population outflows from three counties in Wuhan with many colleges and universities in the first two weeks of January were likely college students leaving the city to avoid the peak traffic just one week before the New Year. Because of the early timing of this movement, many students might have avoided the period when the virus spread rapidly in Wuhan, and their risk of spreading the virus may be low as well. However, our results suggest that during the two weeks prior to Wuhan’s travel ban, a large number of travellers still departed Wuhan into neighbouring cities and other megacities in China and may have spread the virus to new areas, as the timing of the lockdown occurred during the latter stages of peak population numbers leaving Wuhan. Further exacerbating this risk, we found that during the outbreak’s initial stages, a particularly high proportion of cases travelled with illness caused by the virus, together with the transmissibility of 2019-nCoV through asymptomatic contacts, potentially causing additional transmission during travel [28].

Moreover, several destination cities (Figure 5) outside of Hubei Province that received high volume of travellers from Wuhan prior to LNY’s day may serve as significant secondary cities in the outbreak. Most of these cities have large populations and international airports, highly connected with other regions within and beyond China. The initial imported seed cases likely caused the local community transmission, and further spread the virus into wider geographical ranges following the population flows occurring due to the LNY holiday [19]. Therefore, substantial public health interventions have been immediately applied across the country since LNY, including the cordon sanitaire in several of the most affected cities, cancellation of mass gatherings, reduction of travel and contact rate, as well as the extension of the LNY and school winter holiday, which might mitigate subsequent local establishment of 2019-nCoV introduced by travellers.

Beyond the cases that have occurred in China, air passengers have spread 2019-nCoV across countries and continents within a short time period [28, 29]. In particular, a high volume of international airline travellers left Wuhan for hundreds of destination cities across the world during the two weeks prior to the travel restriction implemented in the city. Substantial preparedness efforts at destination cities should be taken to prevent further international seeding and transmission, otherwise the local establishment of epidemics, and even a pandemic, might become inevitable. For example, exit and entry screening may be futher extended to capture travellers with fever who have potentially been exposed to the virus in high-risk Chinese cities where local transmission has been established. However, we estimated that a huge volume of airline travellers would be required to be screened in February to April, even under the scenario of significant reduction (90%) in air passengers compared with the same period of previous years. Therefore, ensuring that surveillance and health systems around the world are ready and sufficiently strong to detect and deal with cases seen is a priority.

It is expected that further international exportation of cases from China will occur and cases may appear in any country [30]. Thus, all countries should be prepared for containment, including active surveillance, early detection, isolation and case management, contact tracing and prevention of onward spread of the 2019-nCoV infection [6, 31]. Of additional concern is that the anticipated destinations of hundreds of thousands of travellers departing China are to low-income or lower-middle income countries, where inadequately resourced medical and public health systems might be unable to detect and adequately manage an imported case of 2019n-CoV, including possible subsequent community spread.

Due to current limited knowledge of the epidemiology of the virus at the time of writing (e.g. the proportion and infectiousness of asymptomatic or subclinical infections) and the rapidly changing situation of the outbreak, the simplicity of our approach to define importation risk can help to quickly update risk assessments, prioritise surveillance, target limited resources and understand the potential of 2019n-CoV introduction at specific destinations [32]. Compared with other studies [33, 34], we explored the various scenarios of travel restriction and used a more comprehensive and spatio-temporally detailed population movement matrix, together with details on the actual final destination cities of air passengers based on the global itinerary dataset. These novel datasets provide new insights on the impacts of internal and international connectivity on potential transmission of this emerging pathogen during the LNY holiday and over the next three months.

Nevertheless, it is important to note that our study has several major limitations. Firstly, while we do present simple scenarios of reduced air travel volumes, our primary analyses assume “business as usual” travel based on previous non-outbreak years, when significant spatio-temporal changes to human travel behaviours across and beyond China have likely occurred recently. Second, the mobile phone data used may provide an incomplete and biased picture of travellers, as the data only cover the population owning a smart phone and using Baidu apps. Third, the case data used in this study likely varies in quality and completeness due to the timeliness of reporting, varying laboratory diagnosis capacities, and differences in details announced on health authority websites. Fourth, compared with airline travellers leaving Wuhan prior to January 23^rd^ evacuees from Wuhan during the January 29^th^ – 31^st^ period might have a higher risk of infection due to their longer stay in Wuhan during the potential continued spread of the virus since January 23^rd^. This may result in overestimates of the number of infections in airline travellers from Wuhan prior to the city’s lockdown. Based on more recent population movement and epidemiological data, we aim to conduct more sophisticated modelling approaches to assess the effectiveness of control measures in China, the impact of movements of people returning from LNY holiday, as well as the risks of a 2019-nCoV global pandemic.

### Ethical statement

Ethical clearance for collecting and using secondary data in this study was granted by the institutional review board of the University of Southampton (No. 48002). All data were supplied and analysed in an anonymous format, without access to personal identifying information.

## Data Availability

The datasets on monthly air passenger data in February – April, 2018 used in this study are available from Dr. Kamran Khan. The case data are available from Dr. Shengjie Lai. The datasets on holidays and air travel statistics from 2010 through 2018 used for validation are available on the WorldPop website (www.worldpop.org). The mobile phone datasets analysed during the current study are not publicly available since this would compromise the agreement with the data provider, but information on the process of requesting access to the data that support the findings of this study are available from Dr. Shengjie Lai.

## Acknowledgments

We thank Baidu Inc. and IATA prodiving the data. We also thank staff members at disease control institutions, hosptials, and health administractions across China where outbreaks occurred for field investigation, administration, and data collection. This study was supported by the grants from the Bill & Melinda Gates Foundation (OPP1134076); the European Union Horizon 2020 (MOOD 874850); the National Natural Science Fund of China (81773498, 71771213, 91846301); National Science and Technology Major Project of China (2016ZX10004222-009); Program of Shanghai Academic/Technology Research Leader (18XD1400300); Hunan Science and Technology Plan Project (2017RS3040, 2018JJ1034). AJT is supported by funding from the Bill & Melinda Gates Foundation (OPP1106427, OPP1032350, OPP1134076, OPP1094793), the Clinton Health Access Initiative, the UK Department for International Development (DFID) and the Wellcome Trust (106866/Z/15/Z, 204613/Z/16/Z). HY is supported by funding from the National Natural Science Fund for Distinguished Young Scholars of China (No. 81525023); Program of Shanghai Academic/Technology Research Leader (No. 18XD1400300); and the United States National Institutes of Health (Comprehensive International Program for Research on AIDS grant U19 AI51915). The research team members were independent from the funding agencies. The funders had no role in the design and conduct of the study; the collection, management, analysis, and interpretation of the data; and the preparation, review, or approval of the manuscript.

## Authors’ contributions

SL: Conceptualised the study, collected data, finalised the analysis, wrote the manuscript draft, revised the manuscript, and interpreted the findings

AJT: Conceptualised the study, revised the manuscript, interpreted the findings

IIB: Data collection and analysis, interpreted the findings and commented on and revised drafts of the manuscript

NWR: Data collection and analysis, interpreted the findings and commented on and revised drafts of the manuscript

AW: Data collection and analysis, interpreted the findings and commented on and revised drafts of the manuscript

XL: Data collection and analysis, interpreted the findings and commented on and revised drafts of the manuscript

The corresponding authors had full access to all the data in the study and had final responsibility for the decision to submit for publication. The corresponding authors attest that all listed authors meet authorship criteria and that no others meeting the criteria have been omitted.

